# Cross-Sectional Validation of an 8-Electrode Multi-Frequency Bioelectrical Impedance Analysis (BIA) Device Against Dual-Energy X-ray Absorptiometry (DEXA) for Body Composition Assessment in Indian Adults

**DOI:** 10.64898/2026.05.24.26353564

**Authors:** Aditya Bheda, Monika Sharma, Neha Jokare, Shreya Kapoor, Jitendra Chouksey

## Abstract

**Background:** Obesity is becoming a global health crisis, and it leads to various metabolic disorders. Body mass index fails to differentiate fat mass from lean mass and systematically misclassifies adiposity risk - a limitation particularly pronounced in South Asian adults, who exhibit characteristically elevated visceral adiposity and reduced appendicular lean mass at a normal BMI. The 2025 Lancet Commission explicitly recommends direct adiposity measurement beyond BMI for obesity diagnosis. Weight loss interventions - whether dietary, behavioural, or pharmacological - are consistently associated with concurrent reductions in both fat mass and lean mass, making body composition monitoring essential beyond scale weight alone. Although DEXA is globally accepted as a gold standard for body composition analysis, the accessibility of DEXA is limited, particularly in resource-constrained low and middle-income countries such as India. BIA devices are a convenient low-cost option to DEXA and can be used for body composition analysis more frequently than a DEXA scan to provide longitudinal data. The aim of this study is to validate 8 electrode BIA devices as a viable alternative to DEXA scan for the South Asian population.

**Methods:** A prospective cross-sectional validation study was conducted following ethics committee approval, with a priori sample size estimation (α = 0.05, power = 80%). Fifty-eight healthy adults (n=58) underwent three BIA measurements and one DEXA scan each. To ensure statistical independence, the three BIA readings per participant were averaged, yielding 58 final measurements for validation. Body fat percentage, lean mass and fat mass were evaluated using Python with statistical analyses like Bland–Altman analysis, Pearson correlation, ICC and regression analysis.

**Results:** In this BIA vs DEXA study, the Pearson correlation was strong across all three outcomes (fat%: r = 0.97; fat mass: r = 0.98; lean mass: r = 0.96), with ICC (2,1) values of 0.94, 0.97, and 0.91 confirming excellent absolute agreement. Mean absolute error was 3.40% for fat percentage, 1.96 kg for fat mass, and 3.37 kg for lean mass. BIA systematically underestimated body fat percentage (bias −1.96%, 95% CI: −2.91% to −1.01%; LoA: −9.04% to +5.12%) and fat mass (bias −0.72 kg, 95% CI: −1.38 to −0.07 kg; LoA: −5.59 to +4.14 kg), while overestimating lean mass by +3.08 kg (95% CI: +2.34 to +3.82 kg; LoA: −2.46 to +8.62 kg).

**Conclusions:** The 8-electrode BIA device shows clinically acceptable agreement with DEXA for body composition assessment in healthy Indian adults. It offers a radiation-free, cost-effective, accessible, and portable alternative to DEXA, making it suitable for longitudinal monitoring and trend detection. The device is particularly valuable for obesity screening and for tracking body composition changes during weight loss interventions at the population level, addressing the critical need for accessible body composition assessment in resource-limited settings.

## 1. INTRODUCTION

The global prevalence of obesity has reached crisis proportions, posing a major public health challenge in the 21st century. A pooled analysis by the NCD Risk Factor Collaboration, covering 3,663 studies and 222 million participants across 200 countries, found that obesity rates more than doubled in adults and quadrupled in children and adolescents between 1990 and 2022 (1). Body mass index (BMI), traditionally calculated as weight in kilograms divided by height in meters squared, has served as the primary metric for obesity classification. However, BMI possesses well-documented limitations: it cannot distinguish between fat mass and lean muscle mass, fails to account for fat distribution patterns, and should not be applied to assess health risk at the individual level (2). In January 2025, the Lancet Diabetes & Endocrinology Commission redefined obesity, introducing ‘clinical’ and ‘preclinical’ obesity. It recommends diagnosing obesity using direct body-fat measures (e.g., DEXA or BIA) alongside BMI to improve risk assessment, as BMI alone is insufficient for individual clinical decisions. This framework, endorsed by over 75 medical organizations, marks a major shift toward body-fat–based diagnosis (3). Body fat percentage (BF%), rather than body weight or BMI alone, has emerged as a clinically meaningful indicator of metabolic health risk. Individuals with a normal BMI but elevated BF% - a condition termed ‘normal-weight obesity’ - face an increased risk of metabolic syndrome and cardiovascular disease (4). While diet-induced weight loss is a cornerstone of obesity therapy, it is accompanied by concomitant loss of lean body mass alongside fat mass, raising concerns for sarcopenia risk; however, global physical function may be preserved or improved owing to reduced fat mass, and mitigation strategies such as adequate protein intake and resistance-type exercise are recommended to attenuate muscle loss (5). These findings collectively reveal a fundamental inadequacy in current clinical practice: while the importance of body composition is increasingly recognised - by regulatory bodies, clinical guidelines, and pharmacological evidence alike - the tools available to measure it accurately, affordably, and repeatedly remain either too imprecise (BMI), too inaccessible (DEXA), or insufficiently validated for the populations that need them most. The question is therefore not whether body composition should be measured, but which tools can do so reliably, and whether they have been validated for the target population. This gap - between the clinical imperative and the methodological reality - defines the rationale for the present study. Among established methods for body composition analysis, DEXA is widely regarded as a gold standard in clinical and research settings (6). However, DEXA’s broader adoption is constrained by practical barriers - including its inaccessibility in resource-limited settings such as India where the obesity burden is rising substantially (7) - and by an inherent methodological limitation: it assumes a constant hydration level, meaning excess body water is measured as additional lean tissue mass and introduces error in body composition estimates (8). A validated, portable, radiation-free alternative deployable without specialist infrastructure is therefore urgently needed.

Bioelectrical impedance analysis (BIA) offers such an alternative - portable, non-invasive, and radiation-free - making it well-suited for routine and longitudinal monitoring (9). Device-specific validation against DEXA in the target population is therefore a prerequisite before BIA measurements can inform clinical or public health decision-making. Three body composition parameters are of particular clinical relevance in the current obesity landscape. First, body fat percentage (BF%) is a direct measure of adiposity recommended by the 2025 Lancet Commission to confirm excess adiposity beyond BMI, and reflects proportional fat content independent of body size (3). Second, fat mass (FM) in absolute kilograms quantifies the actual adipose tissue depot that drives cardiometabolic risk -individuals with a normal BMI but elevated body fat content, a condition termed ‘normal-weight obesity’, face a significantly increased prevalence of cardiometabolic dysregulation, metabolic syndrome, and cardiovascular risk factors (4). Third, lean mass in kilograms - representing skeletal muscle and organ mass - has emerged as a critical monitoring outcome during weight loss. Diet-induced weight loss is consistently associated with concurrent reductions in both fat and lean mass; in persons with overweight or obesity, lean mass typically contributes approximately 20–30% of total weight lost, with the magnitude varying by intervention type and individual characteristics such as sex - underscoring the clinical importance of monitoring body composition beyond scale weight alone (5).

Despite the growing clinical need for accessible and affordable body composition assessment driven by the Lancet Commission on Clinical Obesity’s explicit recommendation for direct adiposity measurement in obesity diagnosis (3), and the expanding emphasis on longitudinal body composition monitoring during weight loss and obesity management - notable geographic and population-specific gaps persist in the literature evaluating modern 8-electrode multi-frequency BIA (MF-BIA) devices against DEXA. Achamrah et al. (10) and Potter et al. (2022) (11) highlight that most BIA–DEXA validation studies have been conducted in White, North American, East Asian, or military populations, with limited data addressing South Asians. Despite the well-described ‘thin-fat’ phenotype of Indian adults - characterised by higher central adiposity and lower lean mass at a given BMI - only a few studies have specifically examined this gap, and the ethnic mismatch likely introduces systematic bias in BIA-derived estimates when applied to Indians. Finally, the Lancet Commission (2025) (3) mandates direct body fat assessment beyond BMI, creating urgent demand for validated, accessible tools in resource-limited settings. The present study was therefore designed to prospectively evaluate the agreement between body fat percentage, lean percentage and lean mass estimated by an 8-electrode multi-frequency BIA device and DEXA-derived measurements in a cohort of 58 healthy Indian adults. The study was approved by an institutional ethics committee.

## 2. METHODOLOGY

### 2.1 Study design and Ethics Approval

This was a prospective, cross-sectional validation study comparing body composition measurements obtained from an 8-electrode multi-frequency BIA device against whole-body DEXA as the reference standard. (6) The study protocol was reviewed and approved by the institutional ethics committee prior to enrolment. All participants provided prior written informed consent.

### 2.2 Study population and sample size

A priori sample size estimation was performed to ensure adequate statistical power to detect agreement and association between BIA and DEXA measurements. An alpha level of 0.05 was used with an assumed statistical power of 80% (12) Based on previous validation studies, a strong expected correlation between methods (r ≈ 0.95) was assumed. Using correlation-based sample size estimation implemented in MedCalc, the minimum required sample size was estimated to be approximately 45–50 (13).

Participants were recruited through an online screening process using a structured questionnaire distributed via social media platforms. Eligibility was assessed based on predefined criteria (provided in the Supplementary data below) including age, sex, and body mass index (BMI).

A total of 66 participants were initially recruited. Data from 58 participants were included in the final analysis, with 8 participants excluded due to protocol non-adherence (e.g., deviations from fasting, timing, or posture requirements). Each participant underwent three BIA measurements and one DEXA measurement under standardized conditions, yielding 174 BIA observations. These data were subsequently averaged for each participant, resulting in a final dataset of 58 independent measurements for statistical validation. Data collection was completed over an 8-week period.

Participants were selected if they were healthy adults aged 18–65 years. Participants were required to be able to stand for at least 2 minutes and lie still for up to 15 minutes to ensure accurate measurement procedures. All assessments were conducted under standardized conditions, including a 12-hour fasting period, abstinence from exercise and alcohol for 24 hours prior to testing, and scheduling of measurements within 1–2 hours of waking in the morning. Participants were instructed to wear light clothing during scans and to remove all metal objects (e.g., zippers, buttons, jewellery) to avoid measurement interference. Individuals were excluded if they had implanted electronic devices or metal implants, were pregnant or breastfeeding, had undergone recent contrast imaging or surgery, had significant fluid imbalance, severe obesity (BMI >40), unstable medical or psychiatric conditions, or any contraindication to low-dose X-ray exposure.

Eligibility and exclusion criteria were based on previously published body composition validation studies, including Holmes *et al.* (8) and Dallman *et al.* (14), as well as established physiological factors affecting bioelectrical impedance and standard manufacturer guidance for DEXA and BIA, to minimize measurement bias and ensure comparability across participants.

### 2.3 Data Collection and Analysis

Data collection was conducted between 12 February and 10 April 2026. Each participant was briefed about the study procedures and provided with legal and informed consent documents to ensure anonymity and uphold the integrity of the research. All BIA and DEXA measurements were performed at Squats Fitness Pvt. Ltd. (FITTR), Kharadi, India, under standardized laboratory conditions. A certified laboratory technician supervised all DEXA scans (GE Healthcare’s Lunar DPX series) to maintain consistency and procedural accuracy. All collected data were anonymized prior to statistical analysis. The dataset included fat percentage, fat mass and lean mass measurements from both the DEXA and BIA methods, along with relevant demographic and anthropometric variables (age, sex, height, weight, and BMI). Initial data were recorded on paper forms, transcribed into Microsoft Excel, and subsequently exported as a Comma Separated Values (CSV) file for analysis using Python 3.12. Data completeness was assessed prior to analysis. No missing values were identified in the final dataset.

### 2.4 Body Composition Measurements

#### 2.4.1 BIA assessment

Body composition was assessed using an 8-electrode multi-frequency BIA device. The device employs an octopolar electrode configuration with two electrodes each at the hand grips and foot platforms, enabling segmental impedance measurement across five body regions (right arm, left arm, trunk, right leg, and left leg). Unlike conventional 4-electrod that estimate whole-body impedance through a single current pathway, the 8-electrode multi-frequency design allows simultaneous segmental current transmission and measurement, improving the resolution of regional body composition estimates, particularly for trunk composition.

Participants stood barefoot on the platform and grasped the hand electrodes in a standardized posture. Measurements were obtained following manufacturer protocols (16).

#### 2.4.2 DEXA assessment

WholeLJbody DEXA scans were performed using a calibrated DEXA equipment. Daily quality assurance calibration was conducted per manufacturer specifications using a calibration block. Participants were scanned in a supine position wearing light clothing without metal objects. Room temperature was controlled within the manufacturer’s specified range (18-27°C). DEXALJderived fat percentage, fat mass and lean mass were used as the reference standard (17).

### 2.5 Statistical analyses

All statistical analyses were performed using Python. Descriptive statistics were reported as mean ± standard deviation (SD), along with minimum and maximum values for DEXA and BIA fat percentage, fat mass, and lean mass values. Error metrics, including mean error (ME), mean absolute error (MAE), standard deviation of differences (SD), and root mean square error (RMSE), were calculated (18).

Agreement between BIA and DEXA was primarily assessed using Bland–Altman analysis. The mean difference (BIA − DEXA) was calculated, and 95% limits of agreement (LoA) were defined as bias ± 1.96 × SD of the differences. Bland–Altman plots were constructed to visualize agreement, including bias and 95% limits of agreement. Statistical significance was set at p < 0.05 (19).

Pearson correlation coefficients (r) and coefficients of determination (r²) were calculated to quantify the strength of linear association. Ordinary least-squares regression was used to derive prediction equations (DEXA = α + β × BIA). Scatter plots were generated to assess the relationship between BIA and DEXA measurements (18).

Intraclass correlation coefficients (ICC) were computed as a supplementary measure of agreement between BIA and DEXA. A two-way mixed-effects model with absolute agreement [ICC (2,1)] was used, as recommended for method comparison studies where one method serves as a fixed reference standard [Koo TK, Li MY. J Chiropr Med. 2016;15(2):155–163]. ICC (3,1) (consistency) was additionally reported (20). Table 1 summarizes the interpretation thresholds for intraclass correlation coefficients (ICC) as proposed by Koo and Li (2016).

**Table 1.** ICC Interpretation Thresholds (Koo & Li, 2016)

| ICC Value Range | Interpretation |
| --- | --- |
| < 0.50 | Poor reliability |
| 0.50 – 0.75 | Moderate reliability |
| 0.75 – 0.90 | Good reliability |
| > 0.90 | Excellent reliability |

Source: Koo TK, Li MY. A Guideline of Selecting and Reporting Intraclass Correlation Coefficients for Reliability Research. J Chiropr Med. 2016;15(2):155–163. doi:10.1016/j.jcm.2016.02.012

Along with this manuscript, the following documents are provided: graphical abstract, raw collected data, consent form, study protocol, investigators brochure, data collection form, ethics committee approval letter and inclusion and exclusion criteria.

## 3. RESULTS

### 3.1 Participant Statistics

A total of 58 participants (32 male, 26 female) were enrolled. Each participant underwent three repeated BIA measurements; values were averaged per participant prior to analysis. Anthropometric characteristics stratified by sex and overall are presented in Table 2 below.

**Table 2:** Anthropometric characteristics of study participants (n=58)

| Parameter | Age (years) | Height (cm) | Weight (kg) | Body Mass Index (kg/m<sup>2</sup>) |
| --- | --- | --- | --- | --- |
| Range | 22 – 61 | 147 – 182 | 48.5 – 99.1 | 18.3 – 36.9 |
| Mean | 38.14 | 165.93 | 70.22 | 25.50 |

### 3.2 Descriptive Statistics

For body fat percentage, the BIA returned a mean BF% of 29.85% ± 10.27% (median: 28.75%; range: 9.00–52.50%), compared to a mean DEXA BF% of 31.81% ± 12.54% (median: 31.30%; range: 8.00–59.00%). Both distributions demonstrated near-symmetrical profiles and were approximately normally distributed.

For fat mass (kg), BIA estimated a mean of 21.31 ± 9.37 kg (median: 19.78 kg; range: 5.43–50.53 kg), compared to a DEXA mean of 22.03 ± 10.91 kg (median: 20.68 kg; range: 5.51–55.00 kg). Both distributions showed mild positive skewness consistent with the right-skewed distribution of adipose tissue in a general healthy adult population.

For lean mass (kg), the BIA estimated a mean of 48.91 ± 9.22 kg (median: 49.03 kg; range: 31.93–69.23 kg), compared to a DEXA mean of 45.83 ± 9.55 kg (median: 45.74 kg; range: 28.88–69.32 kg). Both distributions demonstrated mild positive skewness (BIA: 0.22; DEXA: 0.32), consistent with the right-skewed distribution of lean tissue in a general healthy adult population, and both met normality assumptions.

### 3.3 Error Metrics

For body fat percentage, the mean error (ME, systematic bias) between the BIA and DEXA was −1.96% (95% CI: −2.91% to −1.01%), indicating a statistically significant and consistent underestimation of BF% by the BIA device relative to DEXA. The mean absolute error (MAE) was 3.40%, reflecting the average magnitude of individual measurement discrepancy. The standard deviation of differences (SD) was 3.61%, and the root mean square error (RMSE) was 4.08%.

**Figure 1.**
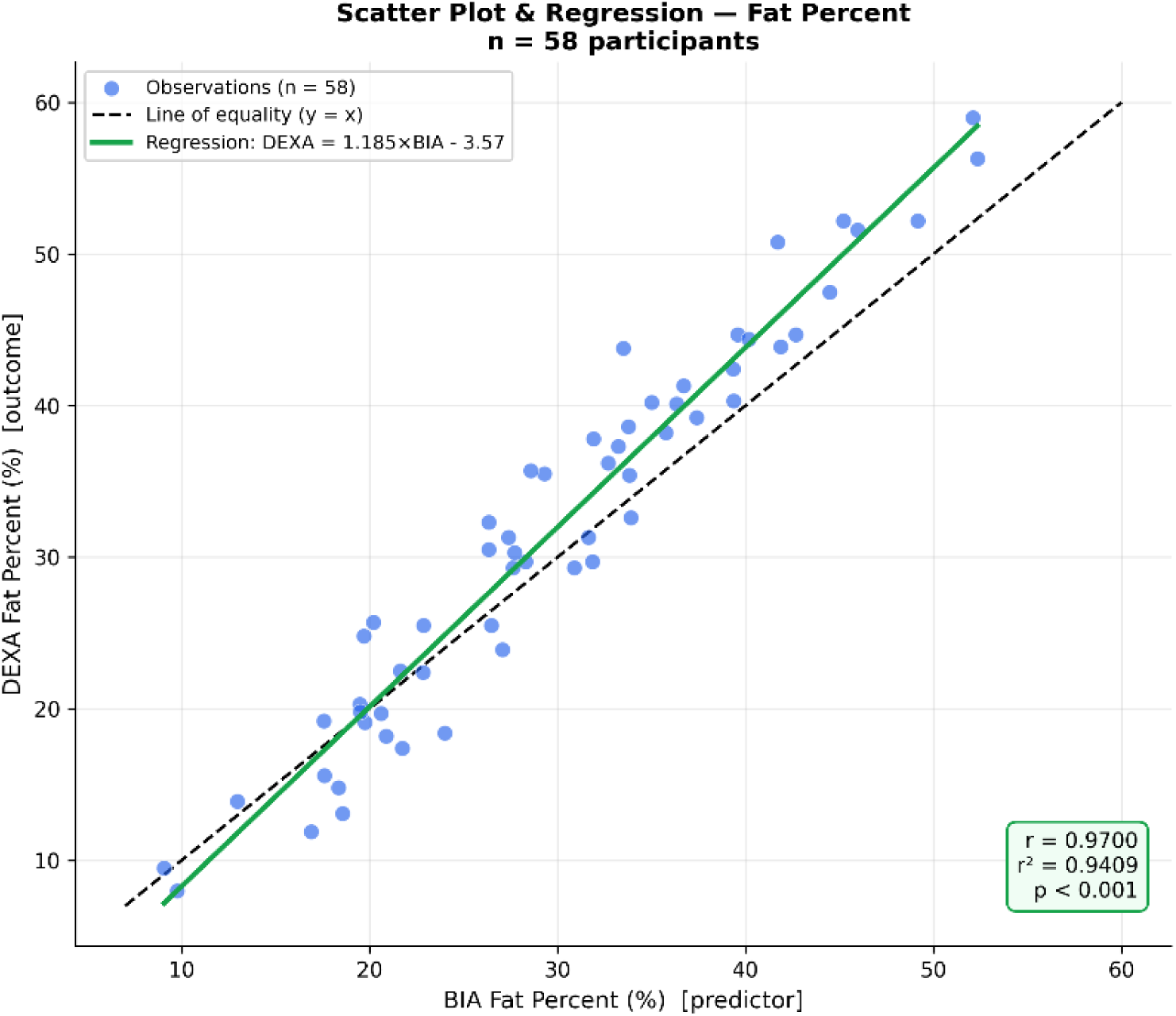
Scatter Plot and Regression Analysis for Fat Percent *Scatter plot showing the relationship between the BIA (x-axis) and DEXA (y-axis) fat percentage measurements (n=58). Each blue dot represents a single averaged measurement. The dashed black line represents the line of equality (y = x), and the solid green line represents the linear regression fit (DEXA = 1.185×BIA − 3.57). The correlation coefficient r = 0.97 with r² = 0.94, indicating that 94.09% of variance in DEXA fat percentage is explained by BIA measurements (p < 0.001). The deviation of the regression line from the equality line indicates systematic bias*.

For fat mass (kg), the ME was −0.72 kg (95% CI: −1.38 to −0.07 kg; p = 0.030), indicating a statistically significant but small systematic underestimation of absolute fat mass by BIA. The MAE was 1.96 kg, SD of differences was 2.48 kg, and RMSE was 2.56 kg. Within-participant agreement was strong: 65.5% of readings fell within ±2 kg and 96.6% within ±5 kg of DEXA values. Cohen’s d was −0.29 (small effect size), confirming the clinical magnitude of this bias is modest.

**Figure 2:**
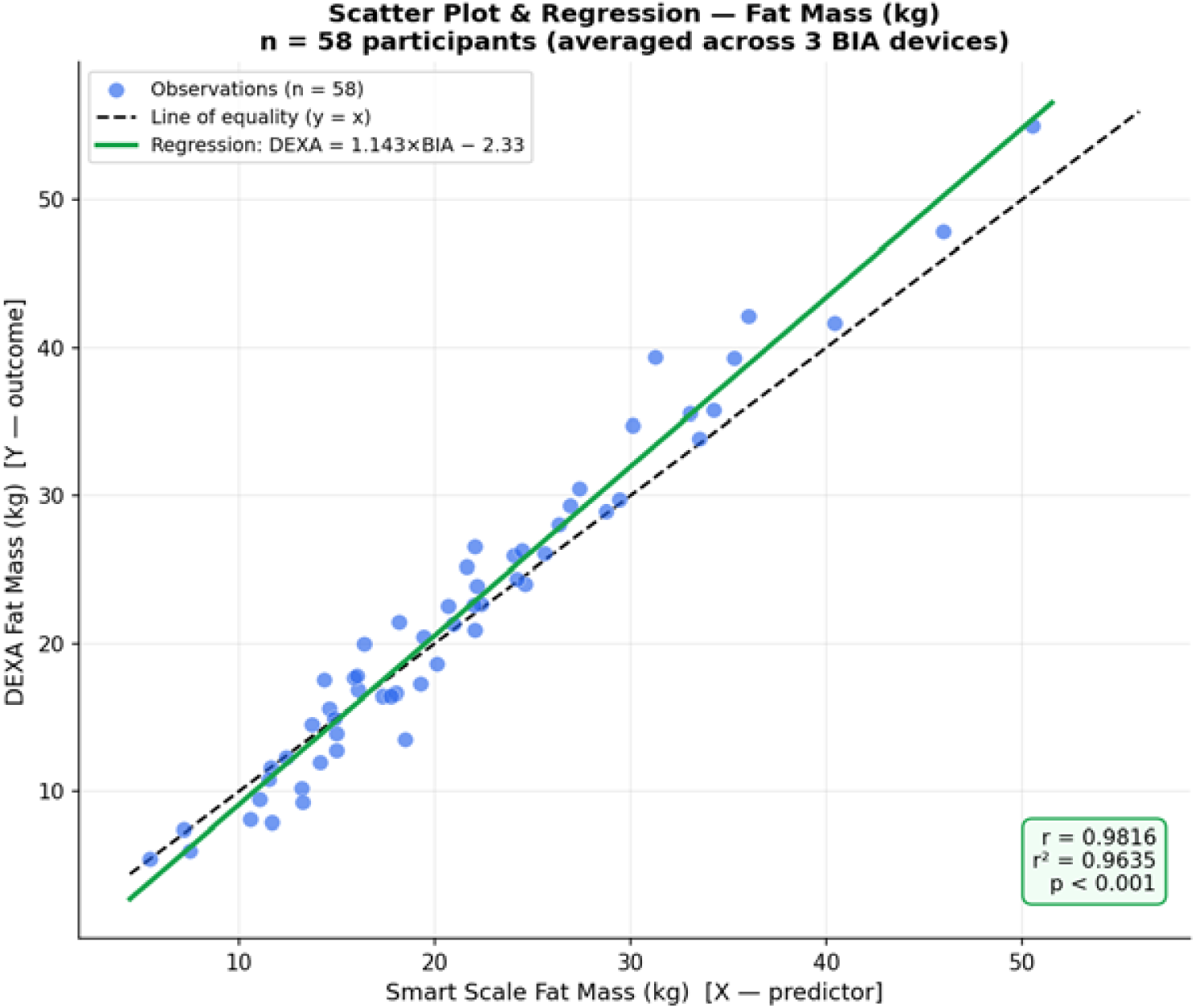
Scatter Plot and Regression Analysis for Fat mass *Scatter Plot and Regression Analysis for Fat Mass (kg) (n = 58). Each blue dot represents a single averaged BIA fat mass measurement plotted against the corresponding DEXA value. The dashed black line represents the line of equality (y = x) and the solid green line represents the linear regression fit (DEXA = 1.143×BIA − 2.333). The correlation coefficient r = 0.98, r² = 0.96, p < 0.001 - the highest correlation observed across all four body composition measures. The close alignment of the regression line with the equality line reflects the small magnitude of systematic bias (−0.72 kg) for fat mass*.

For lean mass (kg), the ME was +3.08 kg (95% CI: +2.34 to +3.82 kg; p < 0.001), indicating a statistically significant and consistent overestimation of lean mass by BIA relative to DEXA. The MAE was 3.37 kg, SD of differences was 2.83 kg, and RMSE was 4.16 kg. Within-participant agreement was lower than that observed for fat mass: only 19.0% of readings fell within ±1 kg and 32.8% within ±2 kg of DEXA values, indicating clinically meaningful individual-level variability despite strong group-level agreement.

**Figure 3:**
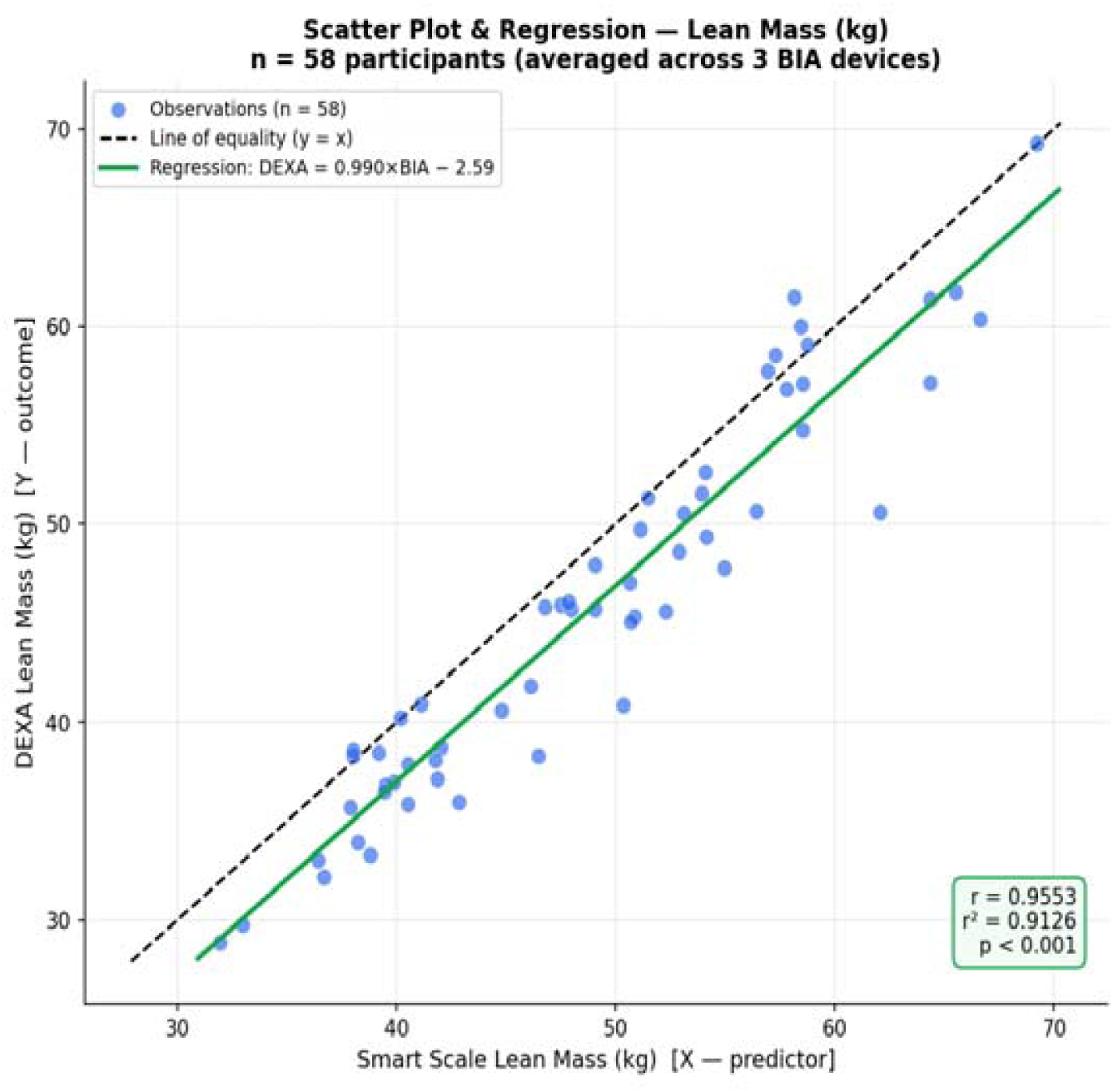
Scatter Plot and Regression Analysis for lean mass *Scatter Plot and Regression Analysis for Lean Mass (kg) (n = 58). Each blue dot represents a single averaged BIA lean mass measurement plotted against the corresponding DEXA value. The dashed black line represents the line of equality (y = x) and the solid green line represents the linear regression fit (DEXA = 0.990 × BIA − 2.585). The correlation coefficient r = 0.96, r² = 0.91, p < 0.001, indicating a strong correlation between methods. The regression line remaining close to the equality line reflects strong proportional agreement across the measurement range, despite the systematic overestimation of lean mass by BIA (+3.08 kg)*.

### 3.4 Correlation and Regression Analysis

Pearson correlation between the BIA and DEXA body fat percentage was r = 0.97 (95% CI: 0.95–0.98; p < 0.001), with a coefficient of determination of r² = 0.94, indicating that 94.00% of the variance in DEXA BF% was explained by the BIA measurements. The regression equation was: DEXA BF% = 1.19 × BIA BF% − 3.57.

Pearson correlation between BIA and DEXA fat mass was r = 0.98 (95% CI: 0.97–0.99; p < 0.001), with r² = 0.96, indicating that 96.00% of the variance in DEXA fat mass was explained by BIA measurements, the highest r² of all four outcomes. The regression equation was: DEXA FM = 1.14 × BIA FM − 2.33. Spearman’s ρ = 0.97 (95% CI: 0.96–0.99; p < 0.001) confirmed that results were robust to distributional assumptions.

Pearson correlation between BIA and DEXA lean mass was r = 0.96 (95% CI: 0.93–0.97; p < 0.001), with r² = 0.91, indicating that 91.30% of the variance in DEXA lean mass was explained by BIA measurements. The regression equation was: DEXA LM = 0.99 × BIA LM − 2.59. The regression slope (0.99), close to unity, indicates that BIA and DEXA scale proportionally across the measurement range despite a fixed absolute offset. Spearman’s ρ = 0.95 (95% CI: 0.91–0.97; p < 0.001) confirmed robustness to distributional assumptions.

### 3.5 Bland-Altman Analysis

Bland-Altman analysis for body fat percentage revealed a mean bias of −1.96% (SD = 3.61%), indicating that the BIA systematically underestimated body fat percentage relative to DEXA. The 95% limits of agreement (LoA) ranged from −9.04% to +5.12% (lower LoA 95% CI: −10.68% to −7.39%; upper LoA 95% CI: +3.48% to +6.77%), representing a total span of 14.16 percentage points.

**Figure 4.**
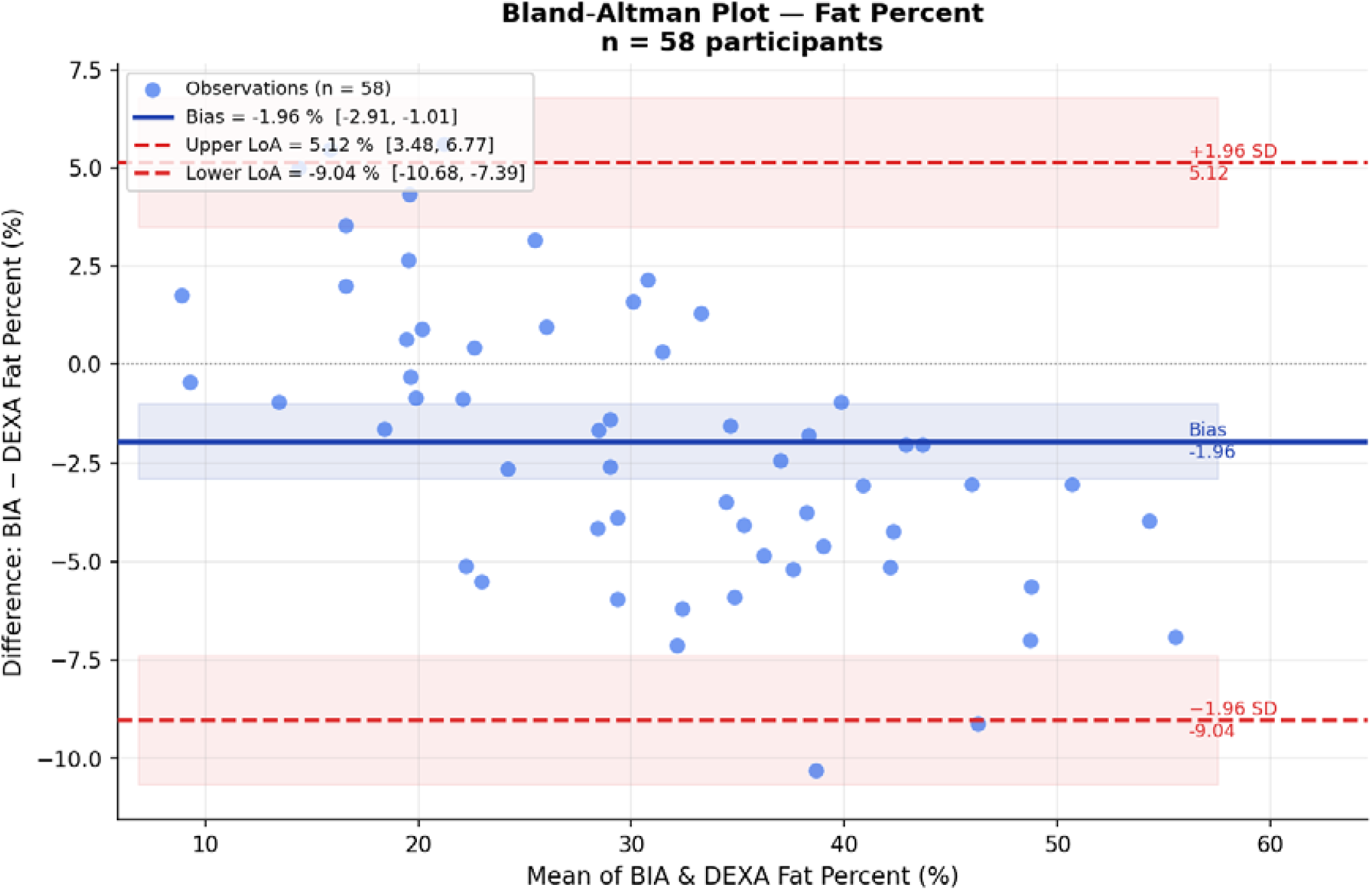
Bland-Altman Plot for Fat Percent Agreement *Bland-Altman analysis comparing the BIA and DEXA fat percentage measurements (n=58). The horizontal blue line represents the mean bias (−1.96%), and the dashed red lines represent the 95% limits of agreement (−9.04% to +5.12%). The shaded region indicates the 95% confidence interval around the bias line*.

Bland–Altman analysis for fat mass revealed a mean bias of −0.72 kg (95% CI: −1.38 to −0.07 kg; SD = 2.48 kg), indicating that BIA systematically underestimated fat mass relative to DEXA. The 95% limits of agreement ranged from −5.59 kg (95% CI: −6.72 to −4.46 kg) to +4.14 kg (95% CI: +3.01 to +5.27 kg), representing a total span of 9.72 kg.

**Figure 5.**
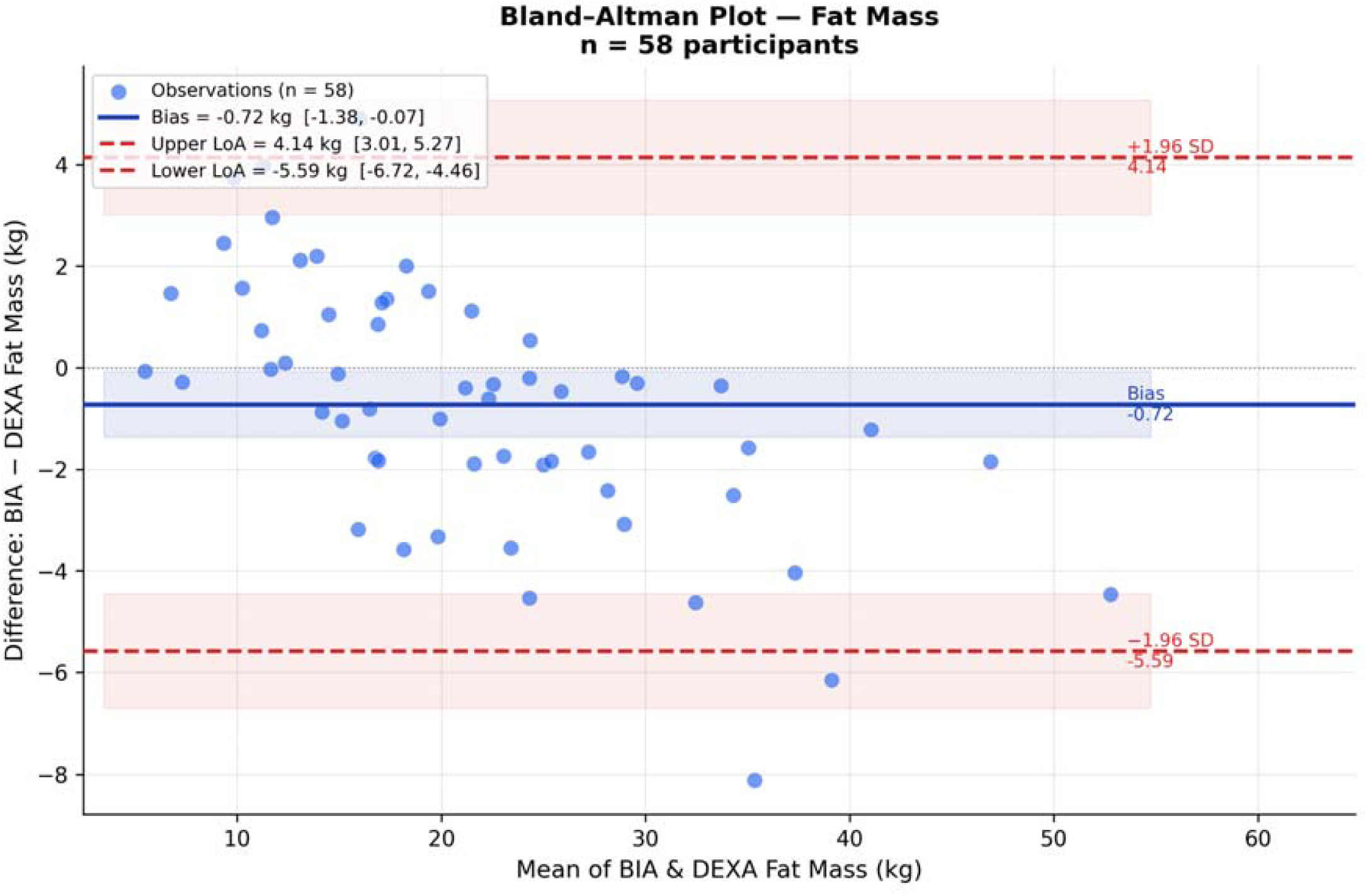
Bland-Altman Plot for Fat mass Agreement *Bland–Altman analysis for lean mass revealed a mean bias of +3.08 kg (95% CI: +2.34 to +3.82 kg; SD = 2.83 kg), indicating that BIA systematically overestimated lean mass relative to DEXA. The 95% limits of agreement ranged from −2.46 kg (95% CI: −3.75 to −1.17 kg) to +8.62 kg (95% CI: +7.33 to +9.91 kg), representing a total span of 11.08 kg*.

Bland–Altman analysis for lean mass revealed a mean bias of +3.08 kg (95% CI: +2.34 to +3.82 kg; SD = 2.83 kg), indicating that BIA systematically overestimated lean mass relative to DEXA. The 95% limits of agreement ranged from −2.46 kg (95% CI: −3.75 to −1.17 kg) to +8.62 kg (95% CI: +7.33 to +9.91 kg), representing a total span of 11.08 kg.

**Figure 6.**
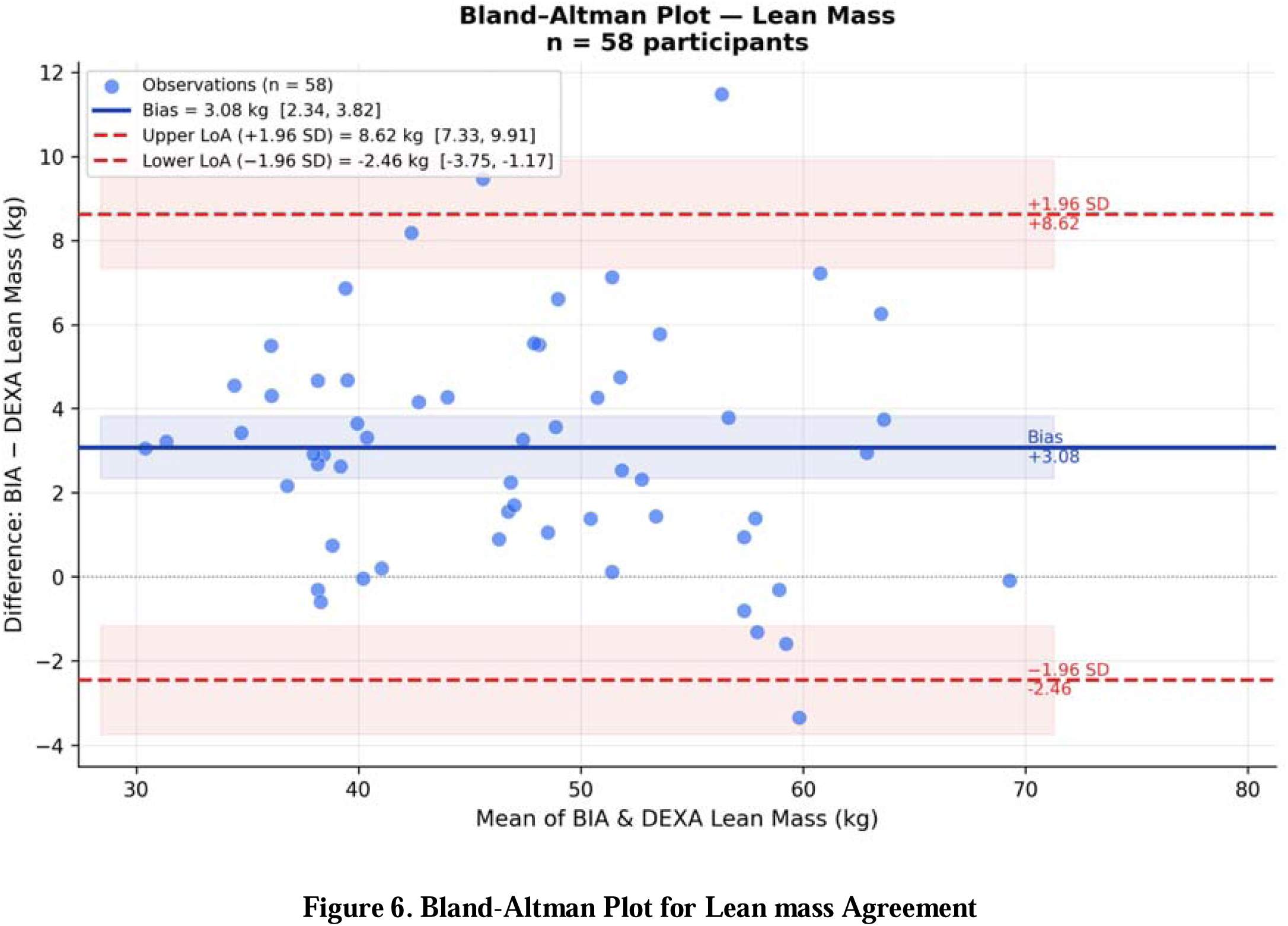
Bland-Altman Plot for Lean mass Agreement *Bland–Altman analysis comparing BIA and DEXA lean mass measurements (n = 58). The horizontal blue line represents the mean bias (+3.08 kg; 95% CI: +2.34 to +3.82 kg). The dashed red lines represent the 95% limits of agreement (−2.46 to +8.62 kg). The shaded region indicates the 95% confidence interval around the bias line*.

### 3.6 Intraclass Correlation Coefficient Analysis

ICC analysis using a two-way mixed-effects model with absolute agreement [ICC(2,1)] demonstrated excellent agreement between BIA and DEXA for both outcomes (Table 3). For fat percentage, ICC(2,1) = 0.94 (95% CI: 0.90–0.96; p < 0.001), indicating excellent absolute agreement. ICC(3,1) for consistency was 0.95 (95% CI: 0.92–0.97; p < 0.001). The difference between ICC(2,1) and ICC(3,1) for fat percentage (0.01) is consistent with the systematic underestimation bias of −1.96% confirmed in the Bland–Altman analysis: the ICC(2,1) is penalised by this fixed offset whereas ICC(3,1) is not.

**Table 3.** Summary of Agreement Metrics Between BIA and DEXA for Body Composition Assessment.

| Metric | Fat Percentage (%) | Fat mass (kg) | Lean Mass (kg) |
| --- | --- | --- | --- |
| DESCRIPTIVE STATISTICS |  |  |  |
| Measurements (n) | 58 | 58 | 58 |
| Mean (BIA device) | 29.85 % | 21.31 kg | 48.91 kg |
| Mean (DEXA) | 31.81 % | 22.03 kg | 45.83 kg |
| ERROR METRICS |  |  |  |
| Mean Bias (95% CI) | -1.96 [-2.91, -1.01] | -0.72 kg [-1.38, -0.07] | +3.08 kg [+2.34, +3.82] |
| MAE | 3.40 % | 1.96 kg | 3.37 kg |
| Standard deviation | 3.61 % | 2.48 kg | 2.83 kg |
| RMSE | 4.08 % | 2.56 kg | 4.16 kg |
| PEARSON CORRELATION |  |  |  |
| Pearson r | 0.97 | 0.98 | 0.96 |
| r<sup>2</sup> | 0.94 | 0.96 | 0.91 |
| 95% CI | [0.95, 0.98] | [0.97, 0.99] | [0.93, 0.97] |
| SPEARMAN CORRELATION |  |  |  |
| Spearman correlation (ρ) | 0.96 | 0.97 | 0.95 |
| 95% CI | [0.94, 0.98] | [0.96, 0.99] | [0.91, 0.97] |
| BLAND- ALTMAN ANALYSIS |  |  |  |
| Upper LoA | +5.12 [3.48, 6.77] | +4.14 [+3.01, +5.27] | +8.62 [+7.33, +9.91] |
| Lower LoA | -9.04 [-10.68, -7.39] | -5.59 [-6.72, -4.46] | -2.46 [-3.75, -1.17] |
| SD of Differences | 3.61% | 2.48 kg | 2.83 kg |
| LINEAR REGRESSION |  |  |  |
| Regression Equation | DEXA = $1.19 \times \text{BIA} - 3.57$ | DEXA = $1.14 \times \text{BIA} - 2.33$ | DEXA = $0.99 \times \text{BIA} - 2.59$ |
| Slope (b) | 1.19 | 1.14 | 0.99 |
| Intercept | -3.57 | -2.33 | -2.59 |
| **Intraclass Correlation Coefficient (ICC)** |  |  |  |
| ICC(2,1) – Absolute Agreement | 0.94 | 0.97 | 0.91 |
| 95% CI | [0.90, 0.96] | [0.95, 0.98] | [0.85, 0.94] |
| p-value | < 0.001 | < 0.001 | < 0.001 |
| ICC(3,1) – Consistency | 0.95 | 0.97 | 0.96 |
| 95% CI | [0.92, 0.97] | [0.95, 0.98] | [0.93, 0.97] |
| p-value | < 0.001 | < 0.001 | < 0.001 |
| ICC(2,1) vs ICC(3,1) | 0.013 (systematic bias effect) | 0.002 (minimal offset) | 0.048 (large bias effect) |

**Notes:** MAE = mean absolute error; RMSE = root mean square error; LoA = limits of agreement; ICC = intraclass correlation coefficient. ICC(2,1): two-way mixed-effects, absolute agreement (recommended for method comparison). ICC(3,1): two-way mixed-effects, consistency. ICC interpreted per Koo & Li (2016): <0.50 poor; 0.50–0.75 moderate; 0.75–0.90 good; >0.90 excellent. Prop. bias slope = regression slope of (BIA – DEXA) on mean of both methods. All analyses n = 58.

ICC analysis for fat mass demonstrated excellent absolute agreement: ICC(2,1) = 0.97 (95% CI: 0.95–0.98; p < 0.001). ICC(3,1) for consistency was 0.97 (95% CI: 0.95–0.98; p < 0.001). The negligible difference between ICC(2,1) and ICC(3,1) (Δ = 0.00) reflects the very small, fixed bias of −0.72 kg, when systematic offset is minimal, absolute agreement and consistency are nearly identical.

ICC analysis for lean mass demonstrated excellent agreement: ICC(2,1) = 0.91 (95% CI: 0.85–0.94; p < 0.001). ICC(3,1) for consistency was 0.96 (95% CI: 0.93–0.97; p < 0.001). The substantial gap between ICC(2,1) and ICC(3,1) (Δ = 0.05) is entirely consistent with the large, fixed bias of +3.08 kg confirmed in the Bland–Altman analysis: ICC(2,1) penalises this systematic offset whereas ICC(3,1) does not. Both values nonetheless exceed the 0.90 threshold for excellent reliability. These findings confirm that BIA tracks lean mass reliably across individuals- the fixed offset is correctable, not variable.

## 4. Discussion and Conclusion

### 4.1 Discussion

#### 4.1.1 Key Findings and Device Performance Against DEXA

This study evaluated agreement between an 8-electrode multi-frequency BIA smart scale and DEXA in 58 healthy Indian adults across three body composition outcomes. For body fat percentage, the device demonstrated strong concordance with DEXA (r = 0.97; r² = 0.94), a mean absolute error of 3.40%, and a systematic underestimation bias of −1.96% (95% CI: −2.91 to −1.01; 95% limits of agreement: −9.04 to +5.12%). For fat mass, concordance was the strongest of all three outcomes (r = 0.98; r² = 0.96), with a mean absolute error of 1.96 kg and a small systematic underestimation bias of −0.72 kg (95% CI: −1.38 to −0.07 kg; 95% limits of agreement: −5.59 to +4.14 kg). For lean mass, the device showed strong correlation (r = 0.96; r² = 0.91), a mean absolute error of 3.37 kg, and a systematic overestimation bias of +3.08 kg (95% CI: +2.34 to +3.82 kg; 95% limits of agreement: −2.46 to +8.62 kg). ICC(2,1) values of 0.94, 0.97, and 0.91 for fat percentage, fat mass, and lean mass, respectively, confirmed excellent absolute agreement across all outcomes.

#### 4.1.2 Systematic Bias and Clinical Implications

The −1.96% mean bias for body fat reflects a well-characterised BIA limitation: algorithms assume fixed relationships between total body water, impedance, and body compartments, which are violated by inter-individual variability in hydration, tissue electrolytes, and body geometry (9,21). Critically, commercial BIA equations are predominantly developed in white and East Asian cohorts; South Asians exhibit distinct phenotypes (higher visceral adiposity, lower skeletal muscle at equivalent BMI) inadequately captured by non-population-specific equations, plausibly accounting for the observed underestimation bias (22). In contrast, lean mass bias was near-zero and statistically non-significant, which is clinically important: the absence of fixed directional lean mass error indicates BIA does not introduce bias into lean mass tracking, supporting its utility for longitudinal monitoring such as detecting lean mass changes during weight loss interventions.

#### 4.1.2 a Statistical Interpretation: Relationship Between Correlation and Agreement

The coexistence of strong Pearson correlation (fat percentage: r = 0.97; fat mass: r = 0.98; lean mass: r = 0.96) and moderate MAE values (3.40%, 1.96 kg, and 3.37 kg, respectively) reflects two distinct but complementary statistical properties. As described in the Bland–Altman framework for method comparison, correlation assesses the strength of linear association between two methods, whereas measures of agreement such as MAE quantify absolute differences at the individual level (19). High correlation does not imply agreement or interchangeability; a perfect correlation may still coexist with substantial systematic bias (19). In addition, intraclass correlation coefficients (ICC) further supported strong agreement between BIA and DEXA, with excellent reliability observed across all components: fat percentage (ICC(2,1) = 0.94, 95% CI: 0.90–0.96; ICC(3,1) = 0.95), fat mass (ICC(2,1) = 0.97, 95% CI: 0.95–0.98; ICC(3,1) = 0.97), and lean mass (ICC(2,1) = 0.91, 95% CI: 0.85–0.94; ICC(3,1) = 0.95), all with p < 0.001.

In this study, r² values of 0.94 (fat percentage), 0.96 (fat mass), and 0.91 (lean mass) indicate that a large proportion of between-individual variability in DEXA-derived measurements was captured by the BIA device, demonstrating strong ranking and tracking capability across participants. In contrast, the MAE values reflect the expected magnitude of deviation when BIA is used as a direct surrogate for DEXA at the individual level. Together, these findings indicate that although the device is not interchangeable with DEXA for single-point diagnostic use, it maintains strong fidelity for relative comparison and longitudinal monitoring of body composition changes (19).

#### 4.1.3 Technological Advantages of 8-Electrode Architecture and Population-Specific Considerations

The 8-electrode (octopolar) BIA system improves upon 4-electrode models by enabling segmental impedance assessment, including direct trunk measurement, thereby enhancing estimation of central adiposity (16). Multi-frequency currents further refine fat-free mass estimation by accounting for intra- and extracellular water distribution. This study provides one of the first prospective validations of an 8-electrode BIA smart scale against DEXA in Indian adults, addressing a key evidence gap. Given the high burden of obesity in India and the distinct ‘thin-fat’ phenotype of South Asians, characterised by higher visceral adiposity at lower BMI, population-specific validation is essential, as existing BIA equations derived from Western and East Asian cohorts may introduce systematic bias when applied to Indian populations (22).

#### 4.1.4 Clinical relevance and comparative value

Intentional weight loss interventions can produce substantial reductions in total body weight, with a meaningful proportion of loss occurring as lean mass, making serial body composition monitoring clinically important (5). In this context, the key value of BIA is not replacement of DEXA for single-point diagnosis, but reliable trend detection across repeated measurements. Although wide limits of agreement limit individual-level interchangeability with DEXA, the near-zero lean mass bias and strong correlation observed here support BIA as a useful tool for longitudinal monitoring without radiation exposure or high cost. Compared with underwater weighing, air-displacement plethysmography, Magnetic Resonance Imaging (MRI), or Computed Tomography (CT) BIA offers a uniquely scalable combination of portability, no radiation, and low cost. This makes it particularly relevant in resource-constrained settings such as India, where access to DEXA is often limited in routine clinical practice, further strengthening the need for scalable alternatives. Rather than matching DEXA exactly, validated BIA can provide sufficiently reliable adiposity signals for screening and follow-up, consistent with broader frameworks that emphasise body fat assessment beyond BMI.

#### 4.1.4a Comparison with Prior 8-Electrode MF-BIA Validation Studies

To contextualise the present findings within the broader BIA validation literature, key accuracy metrics from this study were compared against published validation studies evaluating 8-electrode multi-frequency BIA devices against DEXA as the criterion standard in healthy adult populations (Table 4.1, 4.2, 4.3). Comparator studies were selected based on: (a) use of an octopolar (8-electrode) MF-BIA device; (b) reporting of Bland-Altman agreement statistics; and (c) peer-reviewed publication between 2013 and 2025. Where a specific metric was not reported in the original publication, this is denoted as NR. Notably, several studies demonstrated incomplete reporting of agreement metrics, particularly the absence of mean absolute error (MAE) and, in some cases, coefficient of determination (r²), limiting assessment of individual-level accuracy. In addition, statistical reporting was often non-uniform across body composition compartments, with certain parameters either incompletely evaluated or inconsistently reported. Variability in reported correlation coefficients was also evident across studies; for instance, Sillanpää et al. (2014) reported relatively low correlations for fat mass (r ≈ 0.20) and negative correlations for lean mass (r ≈ −0.30), suggesting potential methodological differences or reduced agreement for specific parameters. Collectively, these factors may constrain the robustness, comparability, and clinical interpretability of findings across studies.

**Table 4.1:** Body Fat Percentage (BF%) Metrics Comparison.

| Study (Year) | Device | n | BF% ME (Bias) | BF% MAE | BF% Pearson r | BF% $r^2$ |
| --- | --- | --- | --- | --- | --- | --- |
| **Present Study (Indian adults, 2026)** | **8-electrode MF-BIA smart scale** | **58** | **-1.96%** | **3.4%** | **0.97** | **0.94** |
| Potter et al., 2022 (BMJ Nutr Prev Health) (11) | InBody 770 (8-electrode MF-BIA) | 795 | -3.4% | 3.9% | NR | 0.96 |
| Looney et al., 2024 (Front. | InBody 770 (8-electrode MF- | 14 | -4.0% | NR | 0.93 | NR |
| Nutr.) (23) | BIA) |  |  |  |  |  |
| Khan et al., 2020 (24) | Omron and InBody 370 | 78 | SF4: -5.5, MF8: -1.1 | NR | SF4: 0.82, MF8: 0.90 | NR |

**Table 4.2:** Body Fat Mass(kg) Metrics Comparison.

| Study (Year) | Device | n | Fat mass ME (Bias) | Fat mass MAE | Fat mass Pearson r | Fat mass r<sup>2</sup> |
| --- | --- | --- | --- | --- | --- | --- |
| **Present Study (Indian adults, 2026)** | **8-electrode MF-BIA smart scale** | **58** | **-0.72 kg** | **1.96** | **0.98** | **0.96** |
| Day et al., 2018 (25) | seca® mBCA 515 | 30 | -0.32 kg | NR | Spearman's rho: 0.98 | NR |
| Sillanpää et al., 2014 (*Obesity, Wiley*) (26) | InBody 720 (8-electrode MF-BIA) | 882 | Women: 3.1 kg; Men: 2.6 kg | NR | Women: 0.201; Men: 0.199 | Men: 0.181; Women: 0.128 |

**Table 4.3:** Body Lean Mass(kg) Metrics Comparison.

| Study (Year) | Device | n | Lean mass ME (Bias) | Lean mass MAE | Lean mass Pearson r | Lean mass r<sup>2</sup> |
| --- | --- | --- | --- | --- | --- | --- |
| **Present Study (Indian adults, 2026)** | **8-electrode MF-BIA smart scale** | **58** | **+3.08 kg** | **3.37 kg** | **0.96** | **0.91** |
| Sillanpää et al., 2014 (*Obesity, Wiley*) (26) | InBody 720 (8-electrode MF-BIA) | 882 | -2.9 kg in women and -1.6 kg in men | NR | Women: r= -0.298<br>Men: r= -0.332 | Women: 0.174; Men: 0.205 |

#### 4.1.5 Limitations, Future Research Directions, and Population Generalisability

Sample-related limitations include the predefined eligibility range of 18–65 years; however, the actual study population comprised individuals aged 22–61 years, thereby underrepresenting younger and older segments within the intended range. Additionally, inclusion was restricted to healthy adults with BMI ≤40, limiting generalisability to individuals with severe obesity, clinical comorbidities, age extremes, or pregnancy. BIA measurements are also sensitive to factors such as hydration status, food and fluid intake, physical activity, ambient temperature, and bladder volume. Although pre-measurement conditions were standardized, adherence to such protocols is not likely to be perfect in real-world settings. Furthermore, single-centre recruitment over a 8-week period may not fully capture the ethnic, regional, and anthropometric diversity of the Indian population.

Priority areas for future research include: (1) extension to clinically important subpopulations (older adults ≥65 years, severe obesity BMI > 40, chronic kidney disease, heart failure, type 2 diabetes); (2) prospective longitudinal validation quantifying device sensitivity for detecting lean mass changes during pharmacological and lifestyle weight loss interventions; (3) development of Indian population-specific BIA prediction equations; (4) multi-centre validation across India’s diverse geographic and dietary contexts to strengthen national obesity screening guidelines aligned with the 2025 Lancet Commission framework.

### 4.2 Conclusion

This study demonstrates that the 8-electrode multi-frequency BIA device achieves strong agreement with DEXA across three body composition outcomes in 58 healthy Indian adults: fat percentage (r = 0.97, bias = −1.96%), fat mass (r = 0.98, bias = −0.72 kg), and lean mass (r = 0.96, bias = +3.08 kg). ICC(2,1) values of 0.94, 0.97, and 0.91, respectively, confirmed excellent absolute agreement across all outcomes. In the context of the 2025 Lancet Diabetes & Endocrinology Commission redefinition of obesity, which calls for direct body fat assessment beyond BMI, and the growing recognition that weight loss interventions are consistently associated with concurrent reductions in lean mass, the need for accessible, affordable, and repeatable body composition monitoring has become increasingly important. Although 8-electrode BIA cannot replace DEXA for individual-level diagnostics because of inherent limits of agreement, it provides a scientifically validated, radiation-free, and cost-effective tool for tracking body composition trends over time. This makes it well suited for monitoring individuals undergoing weight loss interventions, screening under the revised obesity framework, and enabling widespread longitudinal body composition assessment in settings where DEXA is unavailable. Future research should evaluate the sensitivity of BIA to detect clinically meaningful changes in lean mass during pharmacological and lifestyle-based weight management interventions.

## Supporting information

Study Protocol

Consent Form

Inclusion and Exclusion Criteria

Data collection Form

Investigator Brochure

Ethics Committee Approval

Raw Data Version2

## 5. Data Availability

The dataset is provided along with this manuscript.

All study data will be stored securely by Sponsor and Data Fiduciary Squats Fitness Pvt. Ltd. (FITTR), with datAIsm Services Pvt. Ltd. (datAIsm) acting as Collaborator and Data Processor. Data will be anonymized, encrypted in storage and transfer, and retained by Squats Fitness Pvt. Ltd. (FITTR) for at least five (5) years after study completion.

## 6. Disclosure of Generative AI and AI-assisted Technologies

During the preparation of this work the author(s) used AI to assist with drafting of this paper.

## 7. Authors Contribution

A.B. responsible for conceptualization and methodology of the study. A.B and N.J. performed formal statistical analysis and prepared the original draft of the manuscript. A.B. and N.J. contributed to reviewing, editing, and final approval of the manuscript. All authors have reviewed and approved this manuscript.

## 8. Funding

This study was sponsored by Squats Fitness Pvt. Ltd. (FITTR). The study was conducted by datAIsm. The device used for BIA measurement was the Sense Pro. The sponsor had no role in the study design, data collection, analysis, interpretation, or manuscript preparation.

## 9. Conflicts of Interest

The authors declare no conflicts of interest relevant to this manuscript.

## 10. Acknowledgement

This work was supported by Squats Fitness Pvt. Ltd. (FITTR). We gratefully acknowledge the laboratory technicians who performed DEXA scans and maintained quality assurance procedures. We thank the institutional ethics committee for protocol review and approval. We also acknowledge the study participants for their time and cooperation. We acknowledge Squats Fitness Pvt. Ltd. (FITTR) for providing the BIA device and technical specifications.

---

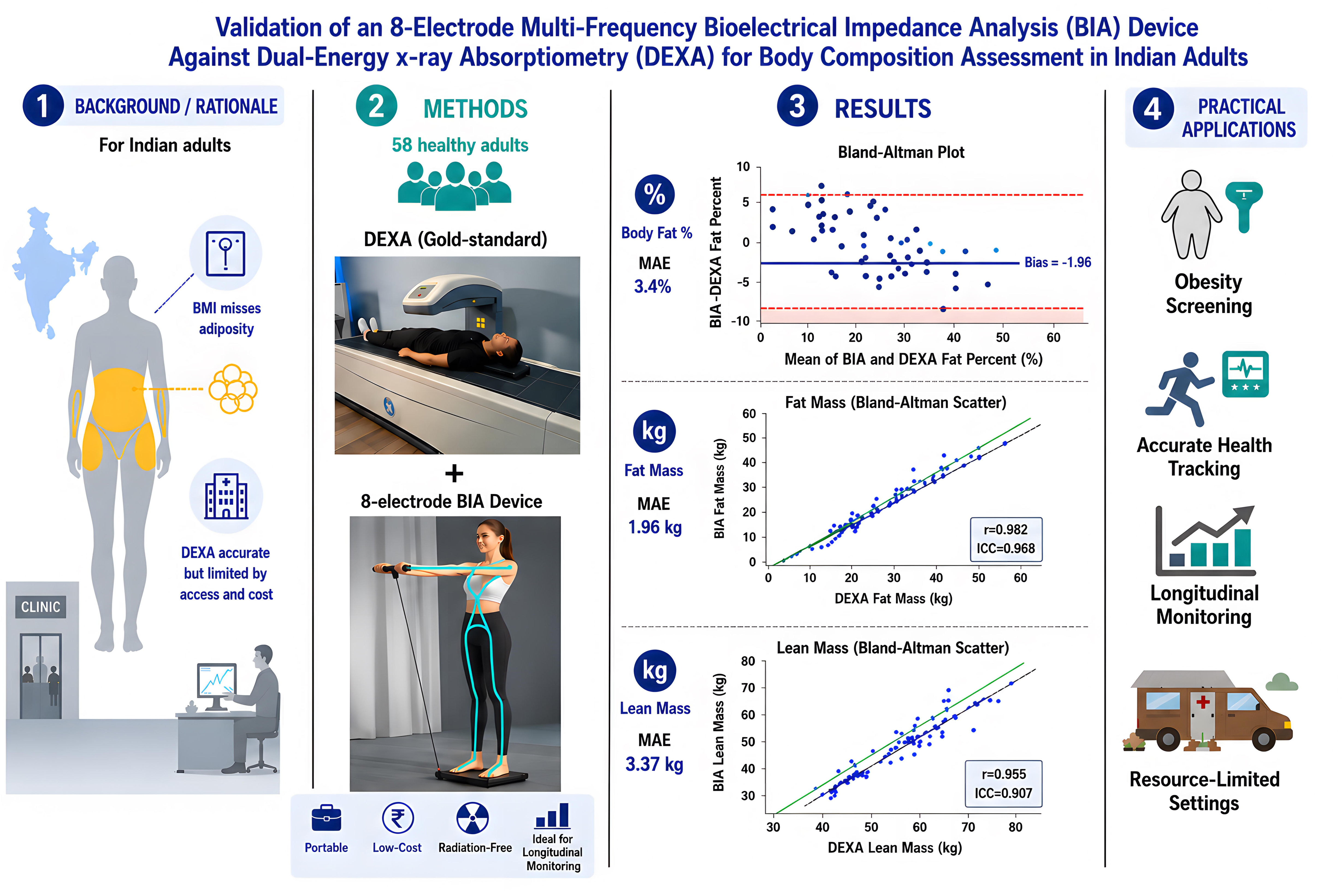

## Notes

### Competing Interest Statement

The authors have declared no competing interest.

### Funding Statement

This study was funded by Squats Fitness Private Limited (FITTR)

### Author Declarations

Supreme Independent Ethics Committee gave ethical approval for this work.

## References

1. Phelps NH, Singleton RK, Zhou B, Heap RA, Mishra A, Bennett JE, et al. Worldwide trends in underweight and obesity from 1990 to 2022: a pooled analysis of 3663 population-representative studies with 222 million children, adolescents, and adults. The Lancet. 2024 Mar;403(10431):1027–50. doi:10.1016/S0140-6736(23)02750-2

2. Nuttall FQ. Body Mass Index: Obesity, BMI, and Health A Critical Review. Nutr Today. 2015 May;50(3):117–28. doi:10.1097/NT.0000000000000092

3. Rubino F, Cummings DE, Eckel RH, Cohen RV, Wilding JPH, Brown WA, et al. Definition and diagnostic criteria of clinical obesity. Lancet Diabetes Endocrinol. 2025 Mar;13(3):221–62. doi:10.1016/S2213-8587(24)00316-4

4. Romero-Corral A, Somers VK, Sierra-Johnson J, Korenfeld Y, Boarin S, Korinek J, et al. Normal weight obesity: a risk factor for cardiometabolic dysregulation and cardiovascular mortality. Eur Heart J. 2010 Mar;31(6):737–46. doi:10.1093/eurheartj/ehp487

5. Cava E, Yeat NC, Mittendorfer B. Preserving Healthy Muscle during Weight Loss. Adv Nutr. 2017 May;8(3):511–9. doi:10.3945/an.116.014506

6. Maeda SS, Peters BSE, Martini LA, Antunes HKM, Gonzalez MC, Arantes HP, et al. Official position of the Brazilian Association of Bone Assessment and Metabolism (ABRASSO) on the evaluation of body composition by densitometry: part I (technical aspects)—general concepts, indications, acquisition, and analysis. Adv Rheumatol. 2022 Dec;62(1):7. doi:10.1186/s42358-022-00241-8

7. Kalra S, Kesavadev J, Goel R, Lakdawala M, Agrawal V, Malhotra N, et al. Burden of Obesity in India: Need for Policy Changes to Attain Highest Possible Level of Health and WellLJBeing. Clin Obes. 2026 Apr;16(2):e70072. doi:10.1111/cob.70072

8. Holmes CJ, Racette SB. The Utility of Body Composition Assessment in Nutrition and Clinical Practice: An Overview of Current Methodology. Nutrients. 2021 Jul 22;13(8):2493. doi:10.3390/nu13082493

9. Dupertuis YM, Jimaja W, Beardsley Levoy C, Genton L. Bioelectrical impedance analysis instruments: how do they differ, what do we need for clinical assessment? Curr Opin Clin Nutr Metab Care. 2025 Sep;28(5):379–87. doi:10.1097/MCO.0000000000001142

10. Achamrah N, Colange G, Delay J, Rimbert A, Folope V, Petit A, et al. Comparison of body composition assessment by DXA and BIA according to the body mass index: A retrospective study on 3655 measures. Handelsman DJ, editor. PLOS ONE. 2018 Jul 12;13(7):e0200465. doi:10.1371/journal.pone.0200465

11. Potter AW, Nindl LJ, Soto LD, Pazmino A, Looney DP, Tharion WJ, et al. High precision but systematic offset in a standing bioelectrical impedance analysis (BIA) compared with dual-energy X-ray absorptiometry (DXA). BMJ Nutr Prev Health. 2022 Dec;5(2):254–62. doi:10.1136/bmjnph-2022-000512

12. Haroun D, Ehsanallah A. Assessment of body fat percentage in Emirati females: a comparative analysis of BIA vs. DXA. Front Nutr. 2026 Jan 9;12:1717492. doi:10.3389/fnut.2025.1717492

13. MedCalc Statistical Software [Internet]. Available from: https://www.medcalc.org/en/calc/

14. Dallman J, Herda A, Cleary CJ, Morey T, Diederich A, Vopat BG, et al. A Brief Review of the Literature for Published Dual-Energy X-Ray Absorptiometry Protocols for Athletes. Sports Health Multidiscip Approach. 2024 Sep;16(5):735–43. doi:10.1177/19417381231208204

15. Von Elm E, Altman DG, Egger M, Pocock SJ, Gøtzsche PC, Vandenbroucke JP, et al. The Strengthening the Reporting of Observational Studies in Epidemiology (STROBE) Statement: Guidelines for Reporting Observational Studies. PLoS Med. 2007 Oct 16;4(10):e296. doi:10.1371/journal.pmed.0040296

16. Bosy-Westphal A, Jensen B, Braun W, Pourhassan M, Gallagher D, Müller MJ. Quantification of whole-body and segmental skeletal muscle mass using phase-sensitive 8-electrode medical bioelectrical impedance devices. Eur J Clin Nutr. 2017 Sep 1;71(9):1061–7. doi:10.1038/ejcn.2017.27

17. Lewiecki EM, Binkley N, Morgan SL, Shuhart CR, Camargos BM, Carey JJ, et al. Best Practices for Dual-Energy X-ray Absorptiometry Measurement and Reporting: International Society for Clinical Densitometry Guidance. J Clin Densitom. 2016 Apr;19(2):127–40. doi:10.1016/j.jocd.2016.03.003

18. Chicco D, Warrens MJ, Jurman G. The coefficient of determination R-squared is more informative than SMAPE, MAE, MAPE, MSE and RMSE in regression analysis evaluation. PeerJ Comput Sci. 2021 Jul 5;7:e623. doi:10.7717/peerj-cs.623

19. Giavarina D. Understanding Bland Altman analysis. Biochem Medica. 2015;25(2):141–51. doi:10.11613/BM.2015.015

20. Koo TK, Li MY. A Guideline of Selecting and Reporting Intraclass Correlation Coefficients for Reliability Research. J Chiropr Med. 2016 Jun;15(2):155–63. doi:10.1016/j.jcm.2016.02.012

21. Kyle U. Bioelectrical impedance analysis?part I: review of principles and methods. Clin Nutr. 2004 Oct;23(5):1226–43. doi:10.1016/j.clnu.2004.06.004

22. Birk N, Kulkarni B, Bhogadi S, Aggarwal A, Walia GK, Gupta V, et al. Machine learning-based equations for improved body composition estimation in Indian adults. Ong E, editor. PLOS Digit Health. 2025 Jun 23;4(6):e0000671. doi:10.1371/journal.pdig.0000671

23. Looney DP, Schafer EA, Chapman CL, Pryor RR, Potter AW, Roberts BM, et al. Reliability, biological variability, and accuracy of multi-frequency bioelectrical impedance analysis for measuring body composition components. Front Nutr. 2024 Dec 3;11:1491931. doi:10.3389/fnut.2024.1491931

24. Khan S, Xanthakos SA, Hornung L, ArceLJClachar C, Siegel R, Kalkwarf HJ. Relative Accuracy of Bioelectrical Impedance Analysis for Assessing Body Composition in Children With Severe Obesity. J Pediatr Gastroenterol Nutr. 2020 Jun;70(6). doi:10.1097/MPG.0000000000002666

25. Day K, Kwok A, Evans A, Mata F, Verdejo-Garcia A, Hart K, et al. Comparison of a Bioelectrical Impedance Device against the Reference Method Dual Energy X-Ray Absorptiometry and Anthropometry for the Evaluation of Body Composition in Adults. Nutrients. 2018 Oct 10;10(10):1469. doi:10.3390/nu10101469

26. Sillanpää E, Cheng S, Häkkinen K, Finni T, Walker S, Pesola A, et al. Body composition in 18LJ to 88 year old adults—comparison of multifrequency bioimpedance and dual energy X ray absorptiometry. Obesity. 2014 Jan;22(1):101–9. doi:10.1002/oby.20583

