## Supplementary material for "Cross-Sectional Validation of an 8-Electrode Multi-Frequency Bioelectrical Impedance Analysis (BIA) Device Against Dual-Energy X-ray Absorptiometry (DEXA) for Body Composition Assessment in Indian Adults": Study Protocol: 3Study Protocol.docx

**PROTOCOL TITLE: Prospective validation study of 8 electrode BIA device for the assessment of key parameters like fat mass, fat percentage, Muscle mass against gold standard Dual-Energy X-ray Absorptiometry (DEXA)**

**SHORT PROTOCOL TITLE:** BIA vs DEXA Study

**PROTOCOL NUMBER:** FRST2503

**VERSION:** 1.0

**VERSION DATE:** 2025-12-03

**STUDY SUMMARY:**

| Investigational Device | 8 electrode BIA smart scale device |
| --- | --- |
| Indicate  Population(s) | Healthy Individuals willing to participate in the study |
| Sample Size | 50+ (progressive data collection) |
| Sponsor | Squats Fitness Private Limited (FITTR) |
| Funding Source | Squats Fitness Private Limited (FITTR) |
| Study Site | Squats Fitness Private Limited (FITTR), Kharadi, Pune |
| Indicate the type of consent to be obtained | ☒Written |
| Research Related | Yes |
| Radiation Exposure | Yes |

### 1 STUDY Objective

The primary objective of this study is to evaluate the accuracy of the 8 electrode BIA smart scale (the BIA device), a smart scale device developed for estimating the body composition, and to quantify its agreement with the standard method of Dual-Energy X-ray Absorptiometry scan (DEXA).

### 2 Background

Body composition assessment is essential for evaluating health status, tracking fitness progress, and managing obesity-related conditions. Traditional methods such as DEXA are considered the practical gold standard for body composition measurement but are expensive, require trained technicians, and expose participants to minimal radiation. Smart scales using bioelectrical impedance analysis (BIA) offer an accessible, cost-effective alternative for home monitoring of body composition. However, the accuracy of these consumer devices requires validation against standards.

DEXA is widely recognized as the preferred standard for quantifying body composition in research settings from a practical standpoint. It provides a three-compartment model (fat mass, lean mass, and bone mineral content) with typical accuracy within 1-3% for body fat percentage. Modern DEXA systems expose participants to very low radiation doses (approximately 0.001-0.01 mSv for a whole-body scan), which is well below natural background radiation levels.

BIA measures the opposition to a small electrical current as it travels through body tissues. While BIA devices demonstrate high reliability (coefficient of variation <2%), their validity compared to DEXA varies considerably. Studies have shown that smart scales can underestimate fat mass by 2-8 kg compared to DEXA, with mean absolute errors (MAE) ranging from 2-6% for body fat percentage.

Validation of the BIA device is necessary to:

- Determine the accuracy of body composition estimates compared to DEXA
- Establish acceptable error margins for consumer applications
- Identify systematic biases and factors affecting measurement accuracy
- Provide evidence-based recommendations for appropriate use

### 3 Study Endpoints

#### 3.1 Primary Study Endpoints

Objective

- To evaluate agreement between the BIA device Fat %, Fat mass, Muscle mass with the standard of DEXA for the same metrics.

Primary Endpoints

Target:

- Achieve MAE ≤ 2.5% for fat % compared to DEXA.
- Achieve MAE ≤ 1.5 kg for fat mass compared to DEXA.
- Achieve MAE ≤ 1.5 kg for fat mass compared to DEXA.

Overall Agreement: Report mean difference (ME) and 95% confidence intervals between the BIA device and DEXA.

Correlation and Agreement: Use Pearson’s r, ICC, and Bland Altman LoA.

- Error Metrics: ME, MAE, RMSE, ErrSTD.
- Classification Accuracy: Fitness level agreement (low, moderate, high).
- Confusion Matrices: Visualize classification accuracy by participant, gender, and BMI.

### 4 STUDY Design and sample size

This is a prospective, cross-sectional, within-subject method comparison study conducted in the FITTR’s diagnostics laboratory. Each participant will undergo a DEXA and then B readings will be taken from 3 separate devices.

Testing will be performed in the DEXA lab with all sessions conducted under consistent air-conditioned temperature. Room temperature was controlled within the manufacturer’s specified range (18-27°C).

The study is designed to evaluate the precision of agreement estimates. A minimum of **50 participants** will provide sufficiently narrow 95% confidence intervals for Bland Altman (BA) agreement statistics and error metrics (MAE, RMSE) when comparing the BIA device with DEXA.

- To account for potential data loss (e.g., poor signal quality, early test termination, device malfunction, or participant withdrawal), additional participants will be recruited.
- The final dataset will ensure **at least 50 complete paired measurements** of the and DEXA are available for analysis.
- Based on literature review the mean error ranges average from 1% to 5% and the standard deviation of error ranges from 2% to 4%. Power analysis with alpha of 0.05, Beta of 0.20, a conservative mean difference of 1.5, and standard deviation of differences of 3.5 yields a minimal required number of pairs as 45. However, it is also expected that different categories of the demography will have different error rates, men vs women, low vs high BMI etc. As we collect more data, this granularity in data will become more reliable- which will in turn benefit in the improvement of the product algorithm after determining the ‘accuracy’ metrics.

### 5 STUDY STAFF ROLES AND RESPONSIBILITIES

1. The Study Data Collection Coordinator (DCC) will collect the data from the subjects before the study and will download and save the study data files obtained from the devices after the study.
2. Study Data Quality Inspector (DQI) will inspect the data collected daily to ensure its validity and will get the errors clarified and updated as required.
3. The Principal Investigator (PI) will lead the entire study, review all the updates daily, and ensure the quality of the study and speed are attained.
4. Technician: Operating Technician will ensure the calibration of the DEXA and make sure the participant is set up for correct study measurement.

### 6 Study Procedures

The following steps will be performed to conduct this validation study.

a) Pre-enrollment (Screening and Setup)

Study participants will need to provide written informed consent to participate in the study after the study's details have been explained to them. Demographic information, including age, sex, height, and weight, medical and physical history, will be collected. Eligible participants will be assigned a unique participant ID, and their data will be recorded.

b) Lab Visit:

At the laboratory visit, a DEXA scan will be done. The BIA device scan will also be performed on the participants using 3 different devices. Investigator’s phone will be used to access the app for the BIA device scan. Participant’s profile will be created in the investigator’s phone. The order of DEXA and smart scale tests will be assigned randomly to reduce the bias. Participants will be instructed to void urine before measurements. Female lab assistant will be present in the lab for female participants.

### 7 Data Collection and Storage

Data collected during the study will be saved in FITTR premises or the FITTR or their vendor’s cloud environment. The data noted on the form will be moved to the data collection excel file by the study coordinator. Regular manual quality audits will be conducted to ensure the collection of high-quality data. If any discrepancies are found, the participant's data will be examined further, and a repeat study will be conducted for the participant. Data will be retained in FITTR's database for at least five years post publication, and periodic audits will ensure compliance and integrity.

### 8 Study Timelines

The study will be completed once quality-checked data from a total of 50 subjects is collected. The study will commence following the success of internal testing, which will involve 50 subjects. This will be followed by a two-week enrollment period, during which 5-10 participants will be recruited per weekday at the FITTR site. Analysis and reporting will be completed in the subsequent week.

The study sample size is subject to expansion as data insights emerge during the study's progression.

### 9  Population Selection and Withdrawal

#### 9.1 Inclusion Criteria

- Participant must be a healthy adult between 18 and 65 years of age
- Participant must be able to stand for 2 minutes
- Participants must be able to lie still for up to 15 minutes.
- Participants should come in for a test 1-2 hours after waking up.
- Participants should fast (12 hours) before both DEXA and smart scale measurements
- Participants should not drink water for 12 hours.
- Participants should not exercise or drink alcohol 24 hours prior.
- Participant Clothing: Wear light clothing, and remove all metal objects for both scans- no metal zippers, metal buttons etc.

#### 9.2 Exclusion Criteria

- Presence of implanted pacemakers, metal implants or other medical devices.
- Severe, unstable medical or psychiatric illness.
- Have any medical condition that could be worsened by exposure to low-dose X-rays.
- Pregnant, suspect you are pregnant or breastfeeding women.
- Recent barium studies or contrast materials (in the last 2 weeks)
- Recent major surgery (in the last 2 weeks)
- Significant fluid retention or oedema (affects BIA accuracy)
- Severe obesity (BMI >40) due to equipment limitations
- Active eating disorders or body dysmorphia
- Any other condition that, in the opinion of the investigator, makes participation unsafe.

### 10 Risks and Benefits

#### 10.1 Risks to Participants

#### Study participation is considered safe, with no significant risks associated with either device. The DEXA scan involves minimal exposure to low-dose X-rays (less than a standard chest X-ray). There are no known risks associated with the smart scale measurement. Some participants may experience mild discomfort from lying still during the DEXA scan. All procedures are supervised to ensure participant safety.

#### 10.2 Potential Benefits to Participants

#### A conveyance allowed of ₹500 will be provided to each participant for this study along with their DEXA Report. The data of the BIA device will not be shared with the participants. This study will help improve the BIA device’s measurement system and do further studies.

### 11 Data Management and Confidentiality

Each participant is assigned a unique identifier, with linkage files stored separately. A data dictionary and case report forms are predefined. All data are stored securely with encryption at rest and in transit, and access is controlled by roles and audited.

### 12 Bias control

Data Engineers conducting preprocessing of study outputs are blinded to subject identifiers.

### 13 Economic Burden to Participants

Subjects will not be subject to any economic burden. They would only contribute their time to the study.

### 14 Ethics and Regulatory

Ethics committee approval will be obtained prior to the start of the study.

### 15 Documentation and audits

Device logs, calibration recordings and study logs will be maintained. Internal audits will be periodically conducted. The study documentation and findings will be stored in FITTR database for any auditing.
