## Supplementary material for "Cross-Sectional Validation of an 8-Electrode Multi-Frequency Bioelectrical Impedance Analysis (BIA) Device Against Dual-Energy X-ray Absorptiometry (DEXA) for Body Composition Assessment in Indian Adults": Consent Form: 4Consent form.docx

**Participant Consent Form for Data Measurement and Collection**

**Study Title:** Prospective Validation Study of FITTR Sense Pro against Dual-Energy X-ray Absorptiometry (DEXA) for Body Composition Assessment.

**Protocol No:** FRST2503

**Study Sponsor (Data Fiduciary):** FITTR Lifecare Pvt. Ltd, 7th Floor, Goodwill B’yond, Kharadi, Pune-411014

**Study Collaborator (Data Processor):** datAIsm Services Private Limited, B2180, Ganga Trueno, Viman Nagar, Pune- 411014, India

**Details of the Investigator:** Aditya Bheda, Consultant, datAIsm Services Private Limited, B2180, Ganga Trueno, Viman Nagar, Pune- 411014, India

**Introduction & Purpose**

This consent form invites participation in a research study titled "Sense Pro Body Assessment Study". The purpose of the study is to compare measurements of fat %, muscle mass, bone mass and other related parameters of the Sense Pro Smart Scale device (Sense Pro) to DEXA clinical equipment. This research aims to determine the accuracy and scientific validity of Sense Pro for body composition assessment. Participation is voluntary, and participants may elect to join or decline without any impact on routine activities.

The study will evaluate whether Sense Pro provides results comparable to a high accuracy device, thereby aiding future research and product development in health monitoring technologies.

**Eligibility Criteria**

- You must be a healthy adult between 18–65 years of age.
- Participants should be able to stand for 2 minutes.
- Participants should be able to lie still for up to 15 minutes.
- Arrive 1–2 hours after waking up in the morning for both DEXA and smart scale measurements.
- You must fast for 12 hours before the test (no food or water).
- You must avoid exercise and alcohol for 24 hours prior.
- Wear light clothing during scans.
- Remove all metal objects (e.g., no metal zippers or buttons) during scans.

You cannot participate if you meet any of the following exclusion criteria:

a. Presence of implanted pacemakers, metal implants or other medical devices.
b. Severe, unstable medical or psychiatric illness.
c. Any medical condition that could be worsened by exposure to low-dose X-rays.
d. Pregnancy, suspected pregnancy, or breastfeeding women.
e. Recent barium studies or contrast materials (in the last 2 weeks)
f. Recent major surgery (in the last 2 weeks)
g. Significant fluid retention or edema (affects BIA accuracy)
h. Severe obesity (BMI >40) due to equipment limitations.
i. Active eating disorders or body dysmorphia.
j. Any other condition that, in the opinion of the investigator, makes participation unsafe.

**Study Procedures**

Those choosing to participate will be assigned a unique identification number and asked to provide demographic details such as age, gender, medical history, and relevant medication use. The procedures involve completing DEXA test which includes lying down on the scan bed as the DEXA equipment scans the body non-invasively from a distance. The second test is an assessment on the Sense Pro which entails standing on the smart scale for 1-2 minutes. Both the tests are performed under direct supervision. Participants may continue normal daily activities outside scheduled test sessions. All personal data and test results will be kept confidential, accessible only to authorized research personnel.

**Data Collection, Usage, and Storage**

All study data will be stored securely by FITTR Lifecare Pvt. Ltd. (Sponsor and Data Fiduciary), with datAIsm Services Pvt. Ltd. acting as Collaborator and Data Processor. Data will be anonymized, encrypted in storage and transfer, and retained by FITTR for at least five (5) years after study completion, in line with protocol FRST2503.

**Risks and Discomforts**

Study participation is considered safe, with no significant risks associated with either device. The DEXA scan involves minimal exposure to low-dose X-rays (less than a standard chest X-ray). There are no known risks associated with the smart scale measurement. Some participants may experience mild discomfort from lying still during the DEXA scan. All procedures are supervised to ensure participant safety.

**Compensation & Benefits**

A conveyance allowed of 500 will be provided to each participant for this study along with their DEXA Report. The data of the study will not be shared with the participant. This study will help improve the BIA device's measurement system also and do further studies.

**Alternatives to Participation**

Participation is voluntary, and individuals may refuse or withdraw at any stage of the research, for any reason, without experiencing any negative consequences or impact on healthcare services.

**Voluntary Participation and Right to Withdraw**

Participants are free to discontinue participation at any time. Withdrawal will not affect access to care or legal rights. If a participant chooses to withdraw, previously collected information remains part of the study analysis, but no new data will be recorded or collected thereafter. All questions and doubts should be addressed before joining.

**Privacy, Confidentiality, and Data Disclosure**

Personal and study data will be strictly protected. Only authorized personnel will have access to the collected information, which will not be shared with third parties except in compliance with legal or ethical requirements. The identity of participants will never be disclosed in any publishable results or official reports.
Any publications or reports arising from this study will use only anonymized data, and no participant will be personally identifiable
FITTR, as the Data Fiduciary, will protect participant rights under the Digital Personal Data Protection Act, 2023. Participants may request access, correction, or deletion of their data by contacting FITTR’s Grievance Officer.

**Contact Information**

Participants who have additional questions, concerns, or requests for information can contact the study investigators or datAIsm representatives using the provided contact details. Support will be available throughout the study duration.

**Acknowledgement and Consent**

By signing below, the participant confirms that the study details, procedures, and potential implications have been explained in clear terms and have been understood. Signing this document signifies voluntary agreement to participate in the Body Assessment study, enabling datAIsm and FITTR to use anonymized data for scientific research purposes. If needed, a legally recognized representative or witness may also sign, ensuring informed and ethical participation.

By signing below, I voluntarily consent to participate in this study and authorize FITTR Lifecare Pvt. Ltd. (Sponsor and Data Fiduciary) and datAIsm Services Pvt. Ltd. (Collaborator/Processor) to process my study data for the purposes described above.

Participant's Name:

Participant’s Signature:

Witness's Name:

Witness's Signature:

Date:
