## Supplementary material for "Cross-Sectional Validation of an 8-Electrode Multi-Frequency Bioelectrical Impedance Analysis (BIA) Device Against Dual-Energy X-ray Absorptiometry (DEXA) for Body Composition Assessment in Indian Adults": Inclusion and Exclusion Criteria: 5Inclusion and Exclusion criteria for the study.docx

Participants were selected based on the predefined inclusion and exclusion criteria provided below:

**Inclusion Criteria**

- Participant must be a healthy adult between 18 and 65 years of age
- Participant must be able to stand for 2 minutes
- Participant must be able to lie still for up to 15 minutes.
- Participant should come in for a test 1-2 hours after waking up.
- Participants should fast (12 hours) before both DEXA and smart scale measurements
- Participants should not drink water for 12 hours.
- Participants should not exercise or drink alcohol 24 hours prior.
- Participant Clothing: Wear light clothing, and remove all metal objects for both scans- no metal zippers, metal buttons etc.

**Exclusion Criteria**

- Presence of implanted pacemakers, metal implants or other medical devices.
- Severe, unstable medical or psychiatric illness.
- Any medical condition that could be worsened by exposure to low-dose X-rays.
- Pregnancy, suspected pregnancy, or breastfeeding women.
- Recent barium studies or contrast materials (in the last 2 weeks)
- Recent major surgery (in the last 2 weeks)
- Significant fluid retention or oedema (affects BIA accuracy)
- Severe obesity (BMI >40) due to equipment limitations.
- Active eating disorders or body dysmorphia.
- Any other condition that, in the opinion of the investigator, makes participation unsafe.
