## Supplementary material for "Cross-Sectional Validation of an 8-Electrode Multi-Frequency Bioelectrical Impedance Analysis (BIA) Device Against Dual-Energy X-ray Absorptiometry (DEXA) for Body Composition Assessment in Indian Adults": Investigator Brochure: 7Investigator Brochure.docx

**Investigator Brochure - BIA Smart Scale Device**

The BIA Smart Scale Device (the BIA device) is a multi‑frequency bioelectrical impedance analysis (MF‑BIA) device designed for quick, non‑invasive assessment of body composition. It provides estimates of body fat percentage, fat mass, lean mass, water content, visceral fat rating, basal metabolic rate, and other derived metrics. The device is intended for fitness and lifestyle tracking rather than medical diagnosis.

**Device Overview**

The BIA device uses an 8‑electrode configuration and applies low‑level electrical currents at multiple frequencies to estimate tissue composition based on impedance patterns. Users input age, height, and gender, after which weight and impedance values are processed to generate outputs. Data syncs automatically via Bluetooth to the companion app, allowing efficient and consistent data capture for studies.

**Specifications**

BATTERY LIFE- 1 charge: 120 days battery life (4.2v Lithium battery)

WEIGHING RANGE- 0.5kg-180kg

COMPATIBILITY- Bluetooth V5.0

MATERIAL- Metallic, Glass

Net weight- 2.7kg

COLOR- Black

**Functions**

Weight, Body fat percentage, BMI, Muscle mass, BMR, Recommended calorie intake, Heart rate, Moisture content, Protein percentage, Fat mass, Lean body mass, Subcutaneous fat percentage, Skeletal muscle percentage, Visceral fat, Bone mass, Muscle percentage, Muscle control quantity, Fat control quantity, Standard weight, Weight control, Body Type, Obesity level, Health assessment, Physical Age, Body Data, Body Score, Skeletal muscle mass, left upper limb fat percentage, Right upper limb fat percentage, Trunk fat percentage, Left lower limb fat percentage, Left upper limb fat mass, Right upper limb fat mass, Trunk fat mass, Left lower limb fat mass, Right lower limb fat mass, Left upper limb muscle mass, Upper limb muscle mass, Right upper limb muscle mass, Trunk muscle mass, Left lower limb muscle mass, Right lower limb muscle mass, Water component, Bone mass, Fat weight, Extracellular water, Cell water, Body cells, Waist-hip ratio, Visceral fat grade, Resting heart rate, Muscle Rate, Body Type, Obesity Level, Health Assessment, Physical Age, Body Score.

**Measurement Protocol**

For consistency, participants should avoid eating or drinking for 2-3 hours, exercising for 12 hours, and alcohol consumption for 24 hours prior to measurement. All readings should be taken barefoot on a hard, level surface, ideally in the morning. Measurements sync automatically through the app, and data can be exported in anonymized form.
