## Supplementary material for "Cross-Sectional Validation of an 8-Electrode Multi-Frequency Bioelectrical Impedance Analysis (BIA) Device Against Dual-Energy X-ray Absorptiometry (DEXA) for Body Composition Assessment in Indian Adults": Ethics Committee Approval: 8Ethics committee Approval.pdf

### Supreme Independent Ethics Committee

HUMAN WELFARE & RIGHTS

**Date:** 18-12-2025

To,
**Dr. Monika Sharma**
Principal Investigator
Squats Fitness Pvt.Ltd (FITTR),
7th Floor, Goodwill B'yond, next to WTC,
Kharadi, Pune, Maharashtra - 411014

**Protocol No: FRST2503**

**Protocol Title:** "Prospective validation study of 8 electrode BIA devices for the assessment of key parameters like fat mass, fat percentage, Muscle mass against gold standard Dual-Energy X-ray Absorptiometry (DEXA)."

**Subject:** Ethics Committee Approval for above mentioned study protocol.

Dear Dr. Monika Sharma,

The Supreme Independent Ethics Committee reviewed and discussed your application to conduct the clinical study mentioned above via online (Google Meet) on 10 December 2025, from 4:15 PM to 4:30 PM.

**A. The following Documents were reviewed:**

| Sr. No | Title | Version and Date |
|---|---|---|
| 1 | Clinical Trial Protocol<br>Protocol no: FRST2503 | Version: 1.0<br>Dated: 03/DEC/2025 |
| 2 | Investigator Brochure | - |
| 3 | Patient Information Sheet/ Informed Consent Form (PIS/ICF) Informed consent form (English) | - |
| 4 | Insurance Policy | 12/11/2025 to 12/11/2026 |

| | | |
|---|---|---|
| 5 | Investigator Undertaking | 03/12/2025 |
| 6 | Investigator CV & MRC | - |
| 7 | Investigator Signature Page | - |
| 8 | electronic Case Report Form (e-CRF) | - |
| 9 | Clinical Trial Agreement (CTA) | 26/11/2025 |

**B. EC MEMBERS**

The Following Members of the Ethics committee were present at the meeting held online via Google Meet on 10 December 2025, from 4:15 PM to 4:30 PM.

| Sr. No | Name | Designation in EC/ Role | Qualification Specialty/Expertise | Gender M/F | Category | Affiliation with the Institute |
|---|---|---|---|---|---|---|
| 1. | Dr. Namdeo Sakharam Jagtap | Chairperson | M.B.B.S, M.D(General Medicine) | M | Scientific | No |
| 2. | Ms. Divya Sharad Borse | Member Secretary | B. Pharm | F | Non-Scientific | Yes |
| 3. | Dr. Namdeo Sakharam Jagtap | Clinician | M.B.B.S, M.D(General Medicine) | M | Scientific | No |
| 4. | Dr.(Mrs) Deepkali Tushar Patil | Clinician | M.B.B.S, M.D (Anesthesiologist) | F | Scientific | No |
| 5. | Dr. Amol Ravindra Hartalkar | Clinician | M.B.B.S, M.D (Medicine) | M | Scientific | No |
| 6. | Dr.(Mrs) Yogita Surendra Karandikar | Basic Medical Scientist | M.B.B.S, M.D(Pharmacology) | F | Scientific | No |
| 7. | Dr. Aman Babanrao Upaganlawar | Basic Medical Scientist | PhD (Pharmacology) | M | Scientific | No |
| 8. | Mr. Gajanan V Karanje | Lay Person | BCS | M | Non-Scientific | No |

| | | | | | | |
|---|---|---|---|---|---|---|
| 9. | Mrs. Vaishali Bapusaheb Awhale | Lay Person | M.S.W(Social Work) | F | Non-Scientific | No |
| 10. | Mrs. Priti Atul Pethkar | Social Scientist | M.S.W(Social Work) | F | Non-Scientific | No |
| 11. | Adv. Swapnil Prakash Pawar | Legal Expert | LLB, LLM | M | Non-Scientific | No |

We approve the trial to be conducted in presented form.

The ethics committee to be informed about the progress of the study every six months. Any Serious Adverse Event (SAE) occurring in the course of the study, any changes in the protocol and patient information or informed consent and to be provided with a copy of the final report.

The study protocol approved with 11 votes in favor of study. It is noted that none of your study members were present during voting for the protocol.

All members of the Ethics Committee confirmed that they had no conflict of interest related to the matters discussed in the meeting,

This Ethics Committee is organized and operates according to the requirements of ICH_GCP, Indian Council of Medical Research (ICMR) and NDCT Rule 2019.

**Note: The approval is valid for two years from the date of approval.**

PSBorse
[illegible]
MEMBER SECRETARY
Supreme Independent Ethics Committee
1856, Bhekarai Nagar, Opposite to IBM Company,
Fursungi, Haveli, Pune-412308.

Thanking You,
Ms. Divya Sharad Borse
**Member Secretary**
1856, Bhekrai Nagar,
Opposite to IBM Company,
Fursungi, Haveli,
Pune Maharashtra -412308 (India)
